# Availability and readiness of healthcare facilities and their effects on long-acting modern contraception use in Bangladesh: Analysis of linked data

**DOI:** 10.1101/2021.09.27.21264197

**Authors:** Md Nuruzzaman Khan, M Mofizul Islam, Shahinoor Akter

**Affiliations:** Department of Population Science, Jatiya Kabi Kazi Nazrul Islam University, Mymensingh-2222, Bangladesh; Department of Public Health, La Trobe University, Melbourne-3086, Australia; Gender and Women’s Health, Centre for Health Equity, School of Population and Global Health, The University of Melbourne, Victoria 3010 Australia

**Keywords:** Modern contraceptives use, contraception, linked data, health facility data, multilevel regression, Bangladesh

## Abstract

**Aim:** Evidence on the availability and accessibility of health facilities and their impacts on long-acting modern contraceptives (LAMC) use in low- and middle-Income countries are scarce. This study examined the influence of the availability and readiness of health facilities in determining the use of LAMC in Bangladesh.

**Methods:** We linked data of the Bangladesh Demographic and Health Survey and the Health Facility Survey using the administrative-boundary linkage method. Mixed effect multilevel logistic regression was conducted. The sample comprised 10,938 married women of 15-49 years of age, who were fertile but did not desire a child within two years of the date of survey. The outcome variable was the current use of LAMC (yes, no) and the explanatory variables were health facility-, individual-, household- and community-level factors.

**Results:** Nearly 34% of participants used LAMC with significant variations across areas in Bangladesh. The average distance between the nearest LAMC-providing health facilities and women’s homes was 6.36 km, higher in the Sylhet division (8.34 km) and lower in the Dhaka division (4.34 km). Increased scores for the management (adjusted odds ratio (AOR) 1.59; 95% CI, 1.21-2.42) and infrastructure (AOR, 1.44; 95% CI, 1.01-1.69) of health facilities were positively associated with the overall uptake of LAMC. AORs for women to report using LMAC were 2.16 (95% CI, 1.18-3.21) and 1.74 (95% CI, 1.15-3.20), respectively, for per unit increase in the availability and readiness scores to provide LAMC at the nearest health facilities. Nearly 27% decline in the likelihood of LAMC uptake was observed for every kilometer increase in the average regional-level distance between women’s homes and the nearest health facilities.

**Conclusion:** The availability of health facilities close to residence and their improved management, infrastructure, and readiness to provide LAMC play a significant role in increasing LAMC uptake among women. Policies and programs should prioritize increasing the availability and accessibility of health facilities that provide LAMC services.

## Introduction

The use of contraception enables individuals or couples to develop their family planning, protect their health^1^ and reduces the risks of maternal morbidity and mortality associated with pregnancy^2^. Contraceptive promotion is, therefore, an integral part of national and international health and population policies, which contributed to a rise in modern contraceptive use from 48% in 1990 to 63% in 2010 worldwide^3^. However, the use of contraception had started declining since then and reached around 50% in 2019^4^. Consequently, around half of the women of reproductive age (15-49 years), 1.1 billion in total, are currently living without modern contraception, although many of them want to use them^4^. About 85 million women of reproductive age, representing 10% of current contraceptive users, use traditional contraceptives ^5^. Over 75% of women with unmet need for contraception live in low- and middle-income countries (LMICs)^6^. Limited access to contraception, inadequate contraceptive options, misinformation, and socio-cultural norms are the predominant factors contributing to the lower use of contraception in LMICs^7^. To revert the unexpected declining trend, global initiatives including the Family planning 2030 initiative ^2^ and Sustainable Development Goals (SDGs) 2030^8^ have emphasised contraception promotion.

Around 44% of all pregnancies in LMICs are unintended at conception, mainly because of the unmet need for contraception. Over 30% of all unintended pregnancies occur due to contraceptive failures, primarily because of using short-term modern contraceptives (e.g., pill and condom) and traditional contraceptives (e.g., withdrawal, rhythm)^9^. In LMICs, unintended pregnancies are responsible for around 55 million unplanned births and 25 million miscarriages in a year^10^. These constitute around 80% burden of pregnancy complications (including severe bleeding and infections)^11^, leading to over 75% of the 118,000 maternal deaths each year in LMICs^7^. Furthermore, such high occurrences of unintended pregnancies in LMICs are attributed to 138 million abortions in a year, including unsafe induced abortion and substantial maternal morbidity and mortality^10^. Due to limited access to long-acting modern contraception (LAMC, the injectable, implant, IUD, female sterilization and male sterillization), about 60 million healthy lives are lost in LMICs per year^12^. Around 10% of child deaths in LMICs are linked with the use of less effective contraceptives, primarily through lower birth intervals^13^ and associated adverse consequences including preterm birth, low birth weight, and small for gestational age^14^.

The SDGs set targets for countries to reduce their maternal mortality ratio (Target 3.1) and ensure universal access to sexual and reproductive health services (Targets 3.7)^8^. To achieve this target, most LMICs, including Bangladesh, are now trying to improve family planning service facilities and the availability of LAMC at those facilities ^5^. However, little is known of the extent to which such endeavour contributes to an increase in LAMC use. This is particularly an issue for Bangladesh and other similar countries where healthcare facilities and family planning services at the community level are functioning under two separate managements and the health system is pularistic^9^. The policymakers of Bangladesh are in a dilemma for over a decade to decide whether the health facility should be given priority over field-level family planning services or merge them^15^. This, to some extent, is due to unclear evidence on how contraception availability in the health facility affects contraception use — a lack that further suppresses the true effects of population-level factors on contraception use ^16-19^.

Increased availability of the geographically referenced health facility and population data creates a window of opportunity to determine the precise effects of health facility-level and population-level factors on LAMC use. However, only a few studies conducted in LMICs^20-23^ have examined these aspects in relation to contraception use in general and LAMC use in particular. Moreover, those studies did not present a stratified analysis of the influence of health facility-level factors on contraception use in rural and urban areas, despite substantial variations in the distribution of health facilities and contraception using patterns in rural and urban areas. This feature is even more prominent in Bangladesh where norms of contraceptive use and associated factors are significantly different across areas^9^. Moreover, studies so far that have been conducted in the context of Bangladesh considered only the population-level factors^9,24,25^. Therefore, the influence of health facility-level factors on LAMC use remains unknown. This study aimed to examine the effects of availability and accessibility of health facility-level factors on LAMC use in Bangladesh.

## Methods

### Study overview

We analysed data extracted from 2017/18 Bangladesh Demographic and Health Survey (BDHS) and 2017 Bangladesh Health Facility Survey (BHFS). We established linkage between the datsets of these two surveys using the administrative boundary linkage method ^26,27^. The Demographic and Health Survey Programme of the USA designed these surveys, and the National Institute of Population Research and Training conducted the surveys under the supervision of the Ministry of Health and Family Welfare of Bangladesh. Detailed sampling procedures of these surveys can be found in the respective survey reports ^28,29^. Briefly, the 2017/18 BDHS collected nationally representative sample data using multistage random sampling methods. At the first stage of sampling, 675 Enumeration Areas (also identified as clusters) were selected from a list of 293579. These clusters had been created as part of the 2011 National Population Census. Data collection was undertaken in the 672 clusters. A household listing operation was conducted and used to select 30 households from each selected cluster through probability proportional to the unit size. Data were collected from 19457 households, with a 96% inclusion rate. There were 20,376 eligible women in the selected households of 15-49 years of age who were usual residents of the selected households or passed there the last night of the day the survey was conducted. Of them, data were collected from 20,127 women with a response rate of 98.8%. The 2017 BHFS used a list of 19,811 registered health facilities as a sampling frame generated by the Ministry of Health and Family Welfare. A total of 1600 health facilities were selected from this list and 1,524 health facilities were finally included in the survey. The selection was made following the census of the district health facilities (DHF) and mother and child welfare centre (MCWC) and stratified random sampling of other healthcare facilities of the government, private, and non-governmental organisations ^29^.

### Sample

We analysed data from 10,384 women selected from 672 clusters included in the BDHS 2017/18. The criteria used for the inclusion in this study were women of reproductive age (15-49), who were fertile but not pregnant or experiencing lactational amenorrhea at the time of the survey and did not desire for a child during two years prior to the date of the data collection.

### Outcome variable

The outcome variable was LAMC use. The relevant data were collected by asking women “*Are you currently doing something or using any method to delay or avoid getting pregnant?*” Women who responded “Yes” to this question were then asked, “*Which method are you using?*” and could select their method from the following list: pill, injectable, implant, intra-uterine device (IUD), condoms, female sterilization, male sterilization, periodic abstinence and withdrawal. A free text option was also provided to report the name of contraception if not included in the list. If a woman reported multiple methods, the method they used most frequently was selected. Responses were coded as “1” for LAMC (i.e., the injectable, implant, IUD, female sterilization and male sterilization) and “0” for contraceptive non-use (responded “no” to the first question) or for use of traditional methods (i.e., periodic abstinence or withdrawal). Women who reported condoms or pills as their current method of contraception were excluded, as these are available at local shops and pharmacies in Bangladesh as well as supplied by the family planning staff working at the field level.

### Explanatory variables

The explanatory variables were identified based on a rapid literature search in the following five databases: PubMed, CINHAL, Web of Science, Embase and Google Scholar. The selected variables were classified into broadly four main groups in line with the multilevel analytic approach: health facility-, individuals-, household-, and community-level factors.

The health facility-level variables were general service readiness (facility’s management and infrastructure), LAMC services availability and readiness scores. For facilities that provide contraception, indices of contraception availability and readiness were created following the WHO guidelines ^30^. Seven variables were used to create contraception availability scores: combined oral contraceptive pills, progestin-only contraceptive pills, progestin-only injectable contraceptives, intrauterine device, implant, male sterilization and female sterilization. These indicator variables were classified in line with this study aim to generate LAMC availability score: given 1 point for each form of LAMC availability in the health facility and 0 for non-availability. Similarly, contraceptive services readiness score was generated using seven dichotomous variables of healthcare personnel availability and training on providing LAMC. These scores were generated using the principal component analysis. The average distance on-road communication of the nearest LAMC-providing facility was also calculated and included as a health facility-level variable. Cluster’s LAMC-providing nearest health facility was identified first. Then using Bangladesh road communication data, the average distance from the cluster to the nearest health facility was calculated, separately for each of the eight administrative divisions. We used regional-level average instead of the actual distance between the clusters and nearest health facilities because the BHFS survey included a sample for all health facilities except DHF and MCWC. Thus, a cluster’s nearest health facility might not have been selected and included in the survey, and hence the actual distance would be problematic. The details of these computation procedures can be found elsewhere ^20^.

The individual-level variables are participants’ age, education, employment status and the number of children ever given birth. The household-level variables are husbands’ education, occupation and household wealth quintile. BDHS generated wealth quintile variable using principal component analysis of the relevant data related to household assets ^31^. The community-level factors were the place of residence (rural and urban) and administrative divisions.

### Statistical analysis

Descriptive statistics were used to summarise the characteristics of the respondents. The global Moran’s I statistic was used to examine the spatial autocorrelation in relation to LAMC use. The Getis-Ord General G statistic was used to measure the degree of clustering of LAMC use. Multilevel logistic regression was used to assess the associations of LAMC use with health facility-, individual- and household-, and community-level factors. The reason for using the multilevel regression was the hierarchical structure of the BDHS data, in which individuals are nested within a household and households are nested within a cluster. This creates multiple dependencies in the data for which multilevel regression is deemed the appropriate approach^32^. We followed the progressive model building technique and developed four different models. Model 1 was the null model where no covariate was adjusted. Model 2 was the health facility-level model where LAMC use was considered with the health facility-level variables. Model 3 was the extension of Model 2 with the inclusion of individual- and household-level factors. All health facility-, individual-, households-, and community-level variables were adjusted in Model 4, which was the final model. We checked multicollinearity before entering these variables into the model. If evidence of multicollinearity was found (VIF>10), the relevant variable was deleted, and the model was run again. Results were reported as Odds Ratios (OR) with 95% Confidence Intervals (95% CI). The Intra-Class Correlation (ICC), Variance Inflation Factor (VIF), Akaike’s Information Criteria (AIC) and Bayesian Information Criteria (BIC) for each model were recorded and compared to select the best model. The ICC value was calculated by dividing the between-clusters-variance of LAMC use (random intercept variance) by the total variance of LAMC use (sum of between-clusters-variance and within-cluster (residual) variance of LAMC use). Statistical package R was used for analyses.

## Results

### Socio-economic characteristics of the respondents

Table 1 shows the background characteristics of the respondents. We analysed data of 10,384 women, 3483 (33.54%) of them used LAMC. The majority of them were in their ages 20-34 years (75%) at the time of the survey. A little more than three-fourths (75.95%) of women had completed primary (36.67%) education, a further one third (32.79%) completed secondary and almost 8% completed further education. Only 4% of women had no children and over 60% had four years or more intervals in their two most recent pregnancies. Around 74% of women resided in rural areas.

**Table 1:**
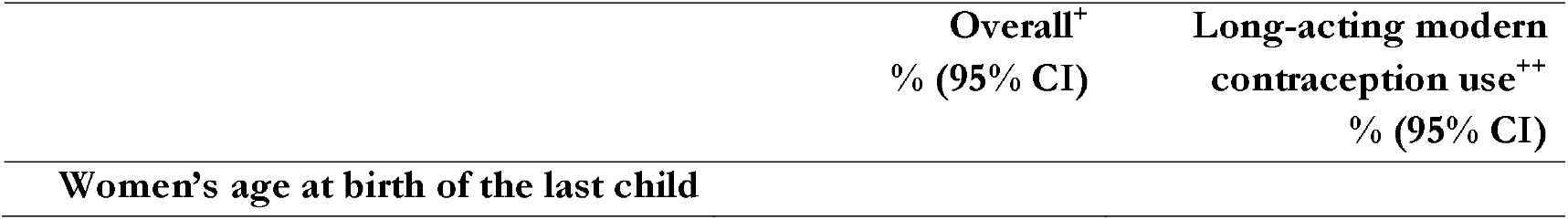
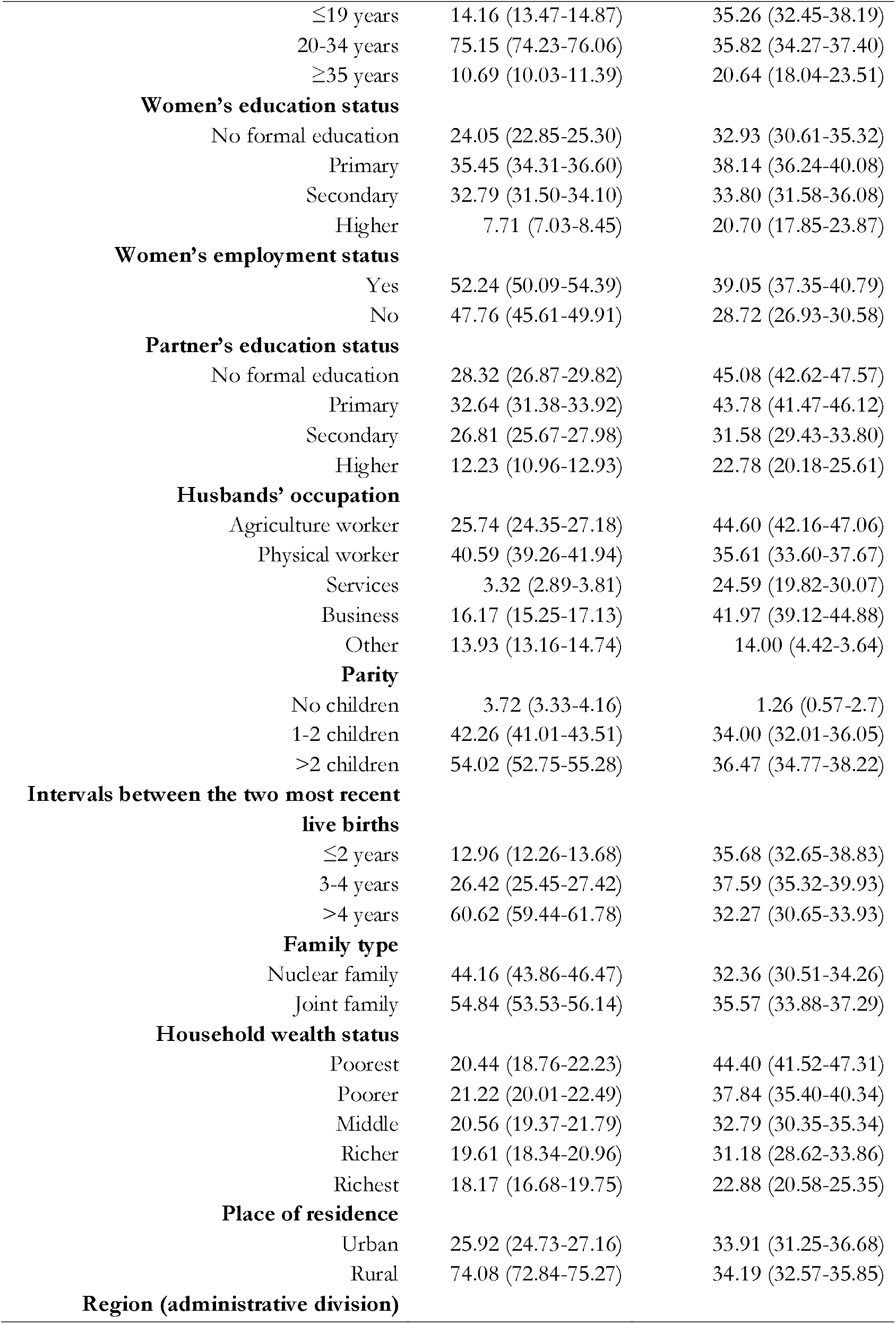
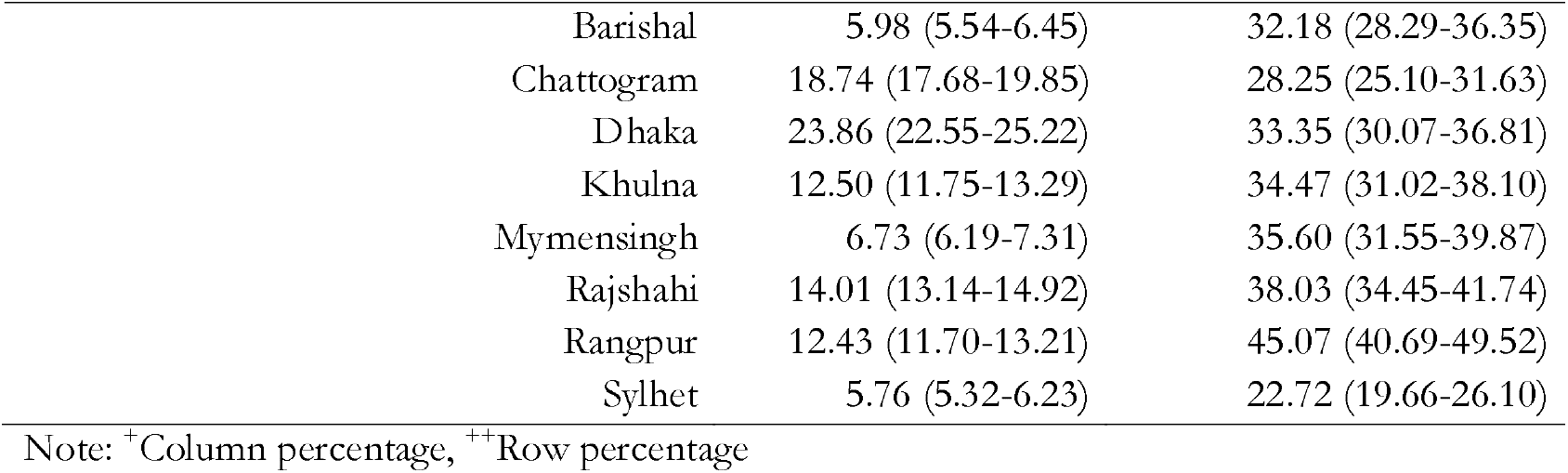
Background characteristics of the respondents, Bangladesh, 2017/18.

|  | Overall<sup>+</sup><br>% (95% CI) | Long-acting modern<br>contraception use<sup>++</sup><br>% (95% CI) |
| --- | --- | --- |
| Women's age at birth of the last child |  |  |

|  |  |  |  |
| --- | --- | --- | --- |
|  | ≤19 years | 14.16 (13.47-14.87) | 35.26 (32.45-38.19) |
|  | 20-34 years | 75.15 (74.23-76.06) | 35.82 (34.27-37.40) |
|  | ≥35 years | 10.69 (10.03-11.39) | 20.64 (18.04-23.51) |
| **Women's education status** |  |  |  |
|  | No formal education | 24.05 (22.85-25.30) | 32.93 (30.61-35.32) |
|  | Primary | 35.45 (34.31-36.60) | 38.14 (36.24-40.08) |
|  | Secondary | 32.79 (31.50-34.10) | 33.80 (31.58-36.08) |
|  | Higher | 7.71 (7.03-8.45) | 20.70 (17.85-23.87) |
| **Women's employment status** |  |  |  |
|  | Yes | 52.24 (50.09-54.39) | 39.05 (37.35-40.79) |
|  | No | 47.76 (45.61-49.91) | 28.72 (26.93-30.58) |
| **Partner's education status** |  |  |  |
|  | No formal education | 28.32 (26.87-29.82) | 45.08 (42.62-47.57) |
|  | Primary | 32.64 (31.38-33.92) | 43.78 (41.47-46.12) |
|  | Secondary | 26.81 (25.67-27.98) | 31.58 (29.43-33.80) |
|  | Higher | 12.23 (10.96-12.93) | 22.78 (20.18-25.61) |
| **Husbands' occupation** |  |  |  |
|  | Agriculture worker | 25.74 (24.35-27.18) | 44.60 (42.16-47.06) |
|  | Physical worker | 40.59 (39.26-41.94) | 35.61 (33.60-37.67) |
|  | Services | 3.32 (2.89-3.81) | 24.59 (19.82-30.07) |
|  | Business | 16.17 (15.25-17.13) | 41.97 (39.12-44.88) |
|  | Other | 13.93 (13.16-14.74) | 14.00 (4.42-3.64) |
| **Parity** |  |  |  |
|  | No children | 3.72 (3.33-4.16) | 1.26 (0.57-2.7) |
|  | 1-2 children | 42.26 (41.01-43.51) | 34.00 (32.01-36.05) |
|  | >2 children | 54.02 (52.75-55.28) | 36.47 (34.77-38.22) |
| **Intervals between the two most recent live births** |  |  |  |
|  | ≤2 years | 12.96 (12.26-13.68) | 35.68 (32.65-38.83) |
|  | 3-4 years | 26.42 (25.45-27.42) | 37.59 (35.32-39.93) |
|  | >4 years | 60.62 (59.44-61.78) | 32.27 (30.65-33.93) |
| **Family type** |  |  |  |
|  | Nuclear family | 44.16 (43.86-46.47) | 32.36 (30.51-34.26) |
|  | Joint family | 54.84 (53.53-56.14) | 35.57 (33.88-37.29) |
| **Household wealth status** |  |  |  |
|  | Poorest | 20.44 (18.76-22.23) | 44.40 (41.52-47.31) |
|  | Poorer | 21.22 (20.01-22.49) | 37.84 (35.40-40.34) |
|  | Middle | 20.56 (19.37-21.79) | 32.79 (30.35-35.34) |
|  | Richer | 19.61 (18.34-20.96) | 31.18 (28.62-33.86) |
|  | Richest | 18.17 (16.68-19.75) | 22.88 (20.58-25.35) |
| **Place of residence** |  |  |  |
|  | Urban | 25.92 (24.73-27.16) | 33.91 (31.25-36.68) |
|  | Rural | 74.08 (72.84-75.27) | 34.19 (32.57-35.85) |
| **Region (administrative division)** |  |  |  |
| Barishal | 5.98 (5.54-6.45) | 32.18 (28.29-36.35) |
| Chattogram | 18.74 (17.68-19.85) | 28.25 (25.10-31.63) |
| Dhaka | 23.86 (22.55-25.22) | 33.35 (30.07-36.81) |
| Khulna | 12.50 (11.75-13.29) | 34.47 (31.02-38.10) |
| Mymensingh | 6.73 (6.19-7.31) | 35.60 (31.55-39.87) |
| Rajshahi | 14.01 (13.14-14.92) | 38.03 (34.45-41.74) |
| Rangpur | 12.43 (11.70-13.21) | 45.07 (40.69-49.52) |
| Sylhet | 5.76 (5.32-6.23) | 22.72 (19.66-26.10) |

Note: <sup>+</sup>Column percentage, <sup>++</sup>Row percentage

### Distribution of long-acting modern contraception use across women’s socio-economic characteristics

We found higher use of LAMC among women aged ≤34 years (36%) compared to those of 35 years or older. Women who received higher education were less likely than others to use LAMC. Women engaged in formal income-generating activities (39%) were more likely than others to report using LAMC. Around a third of women who had a child and 38% of those who had their most recent two births in 3-4 years intervals used LAMC. Higher use of LAMC was found among women in the Rangpur division (45%) and lower among women in the Sylhet division (23%).

### Distribution of health facilities in Bangladesh

The distribution of health facilities across divisions is presented in Table 2. Of the 1524 health facilities, 1357 (89%) provided LAMC and 93% (1262/1357) of these facilities were government hospitals/clinics located at the Upazila or in communities. Relatively high proportions of these facilities were located in the Dhaka (n=245) and Chattogram (n=234) divisions. Sylhet division had a lower number of health facilities with only 87 Upazila/community level governmental hospitals. NGO clinics (n=19) and private hospitals (n=9) were found highly clustered in the Dhaka division. The average distance between the health facilities and the BDHS clusters was 6.36 km, higher in the Sylhet division (8.34 km) and lower in the Dhaka division (4.83 km).

**Table 2:** Division-wise distribution of health facilities that provide long-acting modern contraception and their average distance from the demographic and health survey programme clusters in Bangladesh.

| Division | Availability of long-acting modern contraception (N=1524) |  | Types of health facilities where modern contraception is available (N=1357) |  |  |  |  | Average distance between home and health facility (km) |
| --- | --- | --- | --- | --- | --- | --- | --- | --- |
|  | Yes (n=1357) | No (n=167) | District hospital (n=4) | Upazila to community level government hospital (n=1271) | Mother and Child Welfare Centre (n=7) | NGO clinic or hospital (n=55) | Private hospital (n=21) |  |
| Barishal | 106 (93.71) | 7 (6.29) | 1 (0.36) | 101 (95.88) | 1 (0.65) | 3 (2.73) | 0 (0.00) | 6.43 |
| Chattogram | 261 (90.49) | 27 (9.51) | 1 (0.32) | 245 (93.86) | 1 (0.53) | 9 (3.52) | 5 (1.76) | 5.85 |
| Dhaka | 264 (87.07) | 39 (12.93) | 1 (0.32) | 234 (88.63) | 1 (0.41) | 19 (7.30) | 9 (3.34) | 4.83 |
| Khulna | 175 (93.21) | 13 (6.79) | 1 (0.31) | 165 (94.41) | 1 (0.62) | 7 (4.19) | 1 (0.48) | 5.92 |
| Rajshahi | 201 (91.54) | 19 (8.46) | 0 (0.00) | 191 (94.75) | 1 (0.50) | 7 (3.32) | 3 (1.24) | 7.12 |
| Rangpur | 147 (76.36) | 46 (23.64) | 0 (0.00) | 141 (95.59) | 1 (0.63) | 3 (2.24) | 2 (1.45) | 5.98 |
| Sylhet | 93 (96.13) | 4 (3.87) | 0 (0.00) | 87 (94.22) | 1 (0.50) | 4 (3.54) | 1 (1.41) | 8.34 |
| Mymensingh | 110 (90.0) | 12 (10.0) | 0 (0.00) | 106 (96.43) | 0 (0.00) | 3 (2.66) | 1 (0.88) | 6.44 |
| **Grand average distance** |  |  |  |  |  |  |  | **6.36** |

### Clustering of long-acting modern contraception use in Bangladesh

We found statistically significant positive spatial auto-correlation of LAMC use in Bangladesh (Global Moran’s I=0.374, z=59.486, *p<0*.*001*, results are not shown). The Getis-Ord General G statistics reveals high clustering (z-score= 5.78, *p<0*.*001*). The average distance at which a cluster has at least one neighbour was 202 km. The maximum distance at which clustering of LAMC uses peaked was 674 km. The hot spots, areas with higher LAMC use, were mostly located in the Rangpur and Rajshahi divisions (Figure 1). The cold spots, areas with lower LAMC use, were mostly located in the Sylhet division following the Chattogram, Barishal and Khulna divisions.

**Figure 1:**
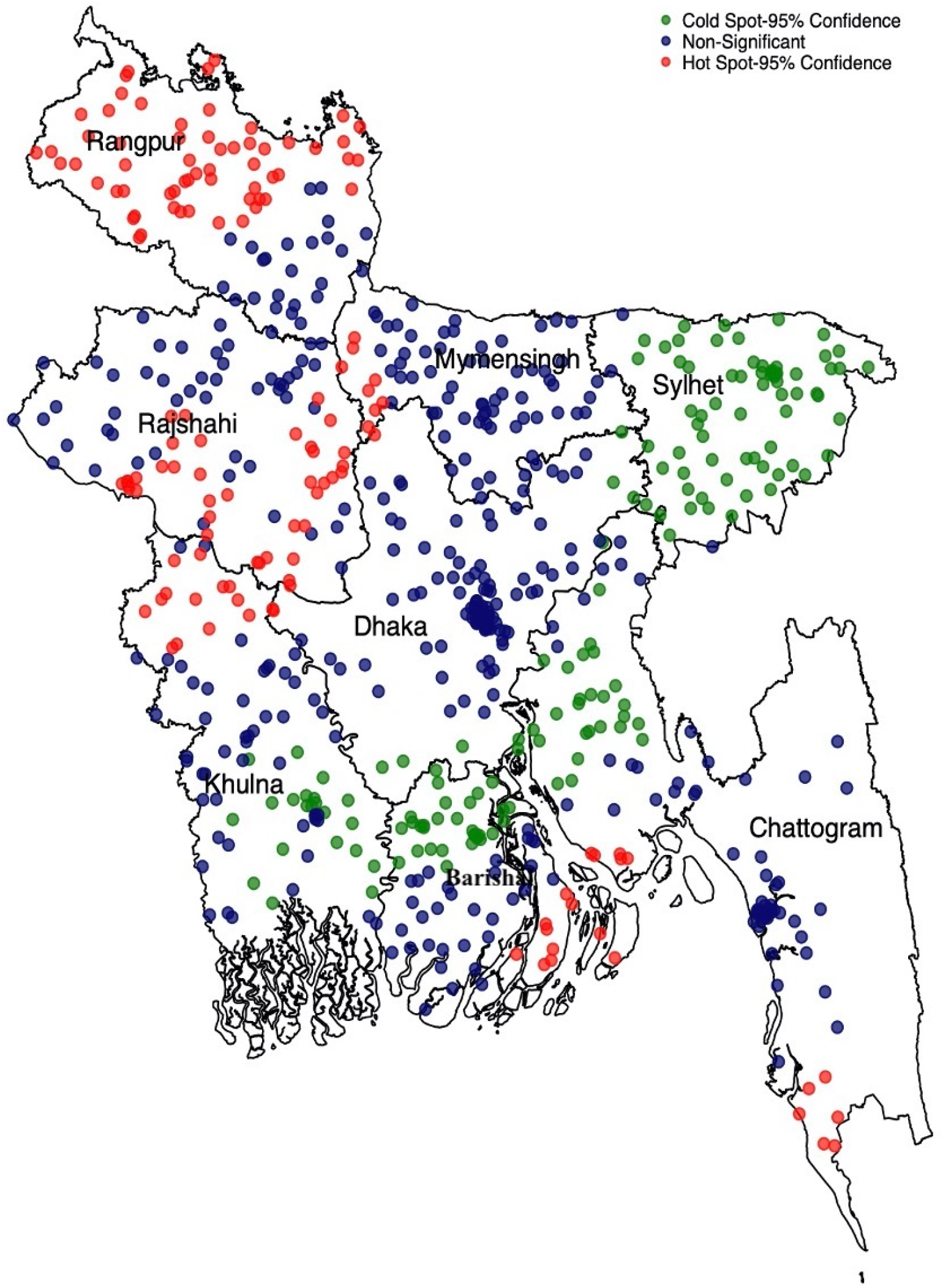
Clusters of high and low use of long-acting modern contraception use in Bangladesh, BDHS 2017/18

### Model selection

The associations of LAMC use with health facility-, individual-, household-, and community-level variables were assessed through multilevel logistic regression models. Of the four models run separately, the AIC, BIC and ICC values were compared to identify the best models. The preferred model was the one that has the smallest AIC, BIC and ICC (Table 3). According to these markers, Model 4 (including health facility-, individual-, household-, and community-variables) fitted the data better. The ICC value for the null model (Model 1) suggested around 17% differences in LAMC use across clusters included in the BDHS. However, this value was reduced to only 3% once health facility-, individual-, household-, and community-level factors were included in the final model. Around 11% of such reduction was reported once health facility-level factors were included in the null model.

**Table 3:** Multilevel logistic regression models assessing the relationships between the use of long-acting modern contraception and health facility-, individual-, household-, and community-level factors (N=10,384)

| Characteristics | Null model | Health facility-level model, OR (95% CI) | Health facility-, individual-, and household-level model, aOR (95% CI) | Health facility-, individual-, household-, and community-level model, aOR (95% CI) |
| --- | --- | --- | --- | --- |
| **General health service readiness** |  |  |  |  |
| Health facility management system |  | 1.76 (1.18-2.36)** | 1.66 (1.28-2.40)** | 1.59 (1.21-2.42)** |
| Health facility infrastructure |  | 1.48 (0.98-1.78) | 1.47 (0.90-1.95) | 1.44 (1.01-1.69)** |
| **The availability of long-acting modern contraception at the health care facility** |  | 2.17 (1.19-3.20)** | 2.23 (1.43-3.76)** | 2.16 (1.18-3.21)** |
| **Readiness of the health care facility to provide long-acting modern contraception** |  | 1.76 (1.14-3.18)** | 1.64 (1.16-3.20)** | 1.74 (1.15-3.20)** |
| **Average distance on road communication from women's resided cluster to the nearest health facility** |  | 0.78 (0.44-0.98)** | 0.74 (0.42-0.97)** | 0.73 (0.46-0.96)** |
| **Women's age** |  |  |  |  |
| ≤19 (ref) |  |  | 1.00 | 1.00 |
| 20-34 |  |  | 0.82 (0.71-0.96)** | 0.84 (0.72-0.97)** |
| ≥35 |  |  | 0.61 (0.48-0.78)** | 0.63 (0.50-0.80)** |
| **Women' education status** |  |  |  |  |
| No formal education (Ref) |  |  | 1.00 | 1.00 |
| Primary |  |  | 1.52 (1.33-1.73)** | 1.55 (1.36-1.77)** |
| Secondary |  |  | 1.73 (1.48-2.03)** | 1.79 (1.53-2.10)** |
| Higher |  |  | 1.33 (1.02-1.74)** | 1.34 (1.03-1.75)** |
| **Women' employment status** |  |  |  |  |
| No (ref) |  |  | 1.00 | 1.00 |
| Yes |  |  | 1.41 (1.27-1.57)** | 1.37 (1.23-1.52)** |
| **Husbands' education status** |  |  |  |  |
| No education (ref) |  |  | 1.00 | 1.00 |
| Primary |  |  | 0.91 (0.81-1.04) | 0.93 (0.82-1.05) |
| Secondary |  |  | 0.60 (0.52-0.69)** | 1.61 (1.53-1.71)** |
| Higher |  |  | 0.43 (0.34-0.55)** | 1.44 (1.34-1.55)** |
| **Husbands' occupation** |  |  |  |  |
| Agricultural worker (ref) |  |  | 1.00 | 1.00 |
| Physical worker |  |  | 0.92 (0.82-1.04) | 0.88 (0.78-0.99)* |
| Services |  |  | 1.24 (0.91-1.90) | 1.18 (0.86-1.61) |
| Business |  |  | 1.29 (1.11-1.48)** | 1.23 (1.06-1.42)** |
| Other |  |  | 0.26 (0.07-0.90)** | 0.26 (0.08-0.91)** |
| **Number of children ever given birth** |  |  |  |  |
| No children (ref) |  |  | 1.00 | 1.00 |
| 1-2 children |  |  | 9.93 (4.70-20.96)** | 9.84 (4.66-20.81)** |
| >2 children |  |  | 9.90 (4.70-20.85)** | 9.92 (4.71-20.91)** |
| **Intervals between the two most recent live births** |  |  |  |  |
| ≤2 years |  |  | 1.00 | 1.00 |
| 3-4 years |  |  | 1.07 (0.91-1.25) | 1.06 (0.90-1.24) |
| >4 years |  |  | 0.97 (0.84-1.13) | 0.95 (0.81-1.10) |
| **Family type** |  |  |  |  |
| Nuclear (ref) |  |  | 1.00 | 1.00 |
| Joint |  |  | 1.18 (1.06-1.30)** | 1.21 (1.10-1.35)** |
| **Household wealth quintile** |  |  |  |  |
| Richest (ref) |  |  | 1.00 | 1.00 |
| Richer |  |  | 1.32 (1.11-1.57)** | 1.41 (1.18-1.68)** |
| Middle |  |  | 1.52 (1.27-1.82)** | 1.71 (1.42-2.06)** |
| Poorer |  |  | 1.81 (1.50-2.18)** | 2.09 (1.72-2.54)** |
| Poorest |  |  | 2.54 (2.09-3.10)** | 2.94 (2.38-3.62)** |
| **Place of residence** |  |  |  |  |
| Urban (ref) |  |  |  | 1.00 |
| Rural |  |  |  | 0.66 (0.57-0.77)** |

| **Region (administrative division)** |  |  |  |  |
| --- | --- | --- | --- | --- |
| Barishal (ref) |  |  |  | 1.00 |
| Chottogram |  |  |  | 1.11 (0.85-1.43) |
| Dhaka |  |  |  | 1.46 (1.13-1.89)** |
| Khulna |  |  |  | 1.18 (0.91-1.53) |
| Mymensingh |  |  |  | 1.16 (0.88-1.53) |
| Rajshahi |  |  |  | 1.44 (1.11-1.87)** |
| Rangpur |  |  |  | 1.59 (1.22-2.06)** |
| Sylhet |  |  |  | 0.77 (0.58-1.02) |
| **Model summary** |  |  |  |  |
| AIC | 12991.51 | 11146.06 | 10969.60 | 10.912.18 |
| BIC | 13006 | 11325.92 | 11139.61 | 11138.87 |
| ICC | 0.17 | 0.11 | 0.10 | 0.07 |
| Median Odds Ratio | 1.80 | 1.78 | 1.78 | 1.68 |
| Variance of the random intercept | 0.38 (0.04) | 0.45 (0.03) | 0.61 (0.04) | 0.55 (0.404) |

Note: \*\* $p < 0.01$ , \* $p < 0.05$

### Factors associated with long-acting modern contraception use

We found health facility-level factors were strong predictors of LAMC use following the adjustment of health facility-, individual-, household-, and community-level factors in the final model. Women were 1.59 times (95% CI, 1.21-2.42) and 1.44 times (95% CI, 1.01-1.69) likely to report using LAMC for every unit increase in the score of health facility management and health facility infrastructure of the nearest health facility, respectively. For each unit increase in the score of contraception service availability at the nearest health facility, the aOR of LAMC use increases to 2.16 (95% CI, 1.18-3.21). Similarly, for every unit increase in the family planning service readiness in the nearest healthcare facility to provide LAMC, the aOR increased to 1.74 (95% CI, 1.15-3.20). The likelihood of women using LAMC reduced by 27% (aOR, 0.73; 95% CI, 0.46-0.96) for every km increase in the average distance on road commucation of health facility from respondents’ resided cluster. At the individual level, lower likelihoods of LAMC use were found among women aged 20-34 years (OR, 0.83; 95% CI, 0.74-0.93) and ≥35 years (OR, 0.56; 95% CI, 0.46-0.67) as compared to the women aged ≤19 years. Women engaged in formal employments and with increased years of education were more likely to report using LAMC than women who received no formal education and were not employed in a formal job.

Women who received primary or more education and whose husbands were received secondary or higher education were significantly more likely than others to use LAMC. Women living in a joint family (aOR, 1.21; 95% CI, 1.10-1.35) were more likely to use LAMC than women living in a nuclear family. LAMC use was around 9.84 times (95% CI, 4.66-20.81) and 9.92 times (95% CI, 4.71-20.91) higher among women with 1-2 children and >2 children, respectively, compared to women who had no children. A gradient of increased likelihoods of LAMC use was found among women with an increasing level of household wealth.

The likelihood of LAMC use was around 41% lower (95% CI, 0.61-0.77) among rural women compared to urban women. Women in the Dhaka (OR, 1.46; 95% CI, 1.13-1.89), Rajshahi (OR, 1.44; 95% CI, 1.11-1.87), and Rangpur (OR, 1.59; 95% CI, 1.22-2.06) divisions were more likely to use LAMC than the women in the Barishal division.

### Effects of health facility environment on long-acting modern contraception use

We calculated the health facility environment by considering all health facilities providing LAMC in 6.3 km (division level average distance on road-communication of health facility from the BDHS cluster) buffer distance. The number of health facilities and their characteristics were generated for the overall and urban and rural locations (Table 4). The likelihood of LAMC uses increased with the increase in the number of health facilities within 6.3 km (aOR 2.01; 95% CI 1.67-2.37). The likelihood increased further (aOR 3.39; 95% CI 1.84-4.90)) if the 6.3 km buffer distance had more than one health facility compared to having no health facility. These likelihoods increased even further for rural areas. The general health services readiness, contraception service availability and readiness were significant predictors of LAMC use.

**Table 4:**
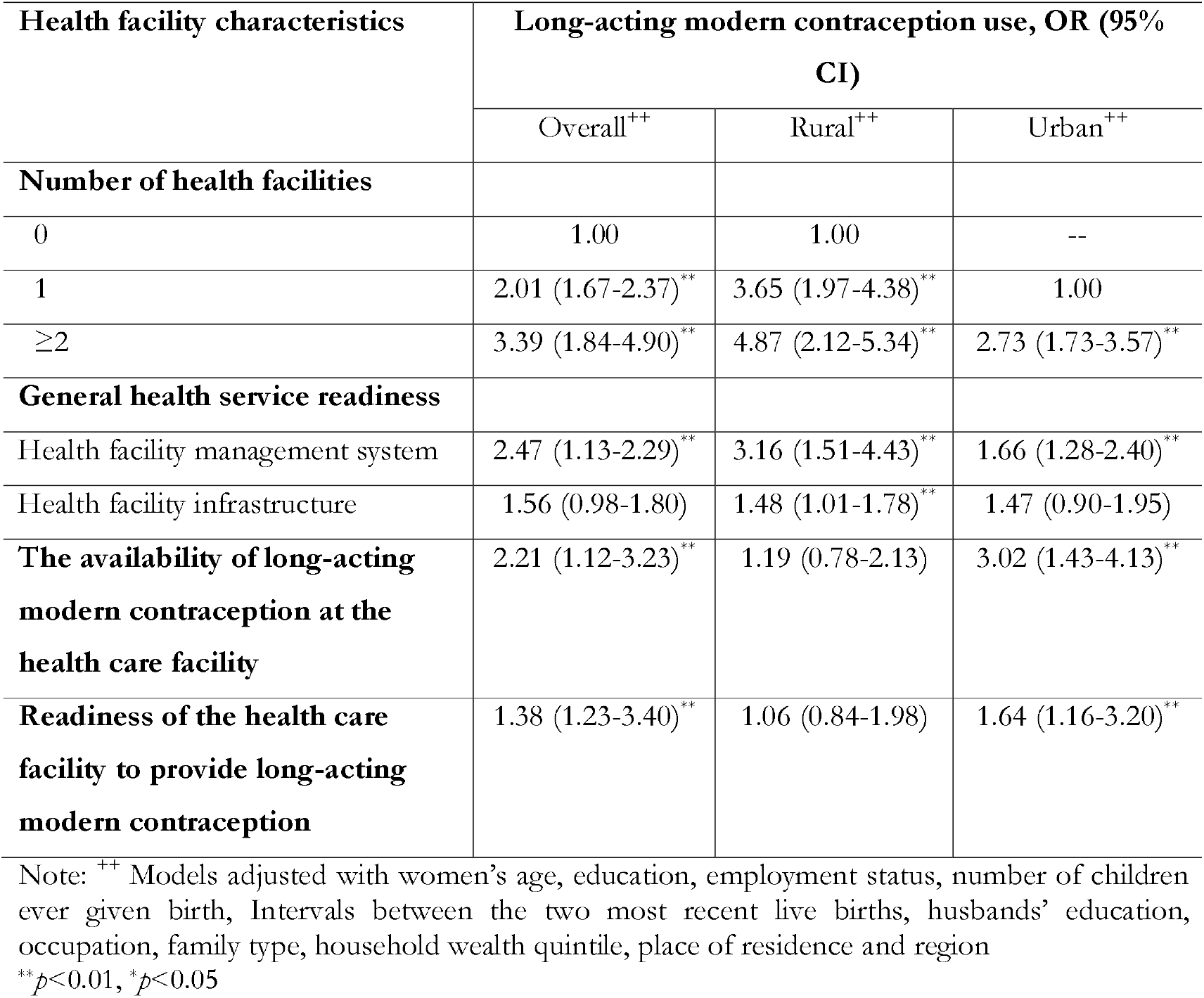
Multilevel logistics regression assessing the relationships between of LAMC use and attributes of health facilities located within 6.36 km distance from the BDHS clusters.

## Discussion

This study provides new insights for LMICs and Bangladesh on the role of availability and accessibility of health facilities in determining LAMC use. Only 33.54% of the total respondents analysed used LAMC. LAMC methods are available in 89% of the health facilities in Bangladesh, over 93% of them were government-owned health facilities. The findings suggest the average distance of a health facility that provides LAMC from the respondents’ home was 6.36 km, highest in Sylhet division (8.43 km) and lowest in Dhaka division (4.34 km). The LAMC uptake was positively associated with LAMC availability at the nearest health facility and its improved management and infrastructure. These relationships were stronger in rural than urban areas. Moreover, the number of health facilities providing LAMC within 6.3 km was revealed as the strongest predictor of LAMC use. Women from lower socio-economic backgrounds were more likely than women of other wealth groups to use LAMC. Overall, our findings suggest that LAMC uptake would increase among women if there are available LAMC-providing facilities with good infrastructure in proximity to women’s homes.

Although the SDGs target to ensure universal coverage of sexual and reproductive healthcare services by 2030^33^, the Bangladesh government has set the target to reach 75% coverage of modern contraception use by 2025^34,35^. Clearly, achieving this target is a challenge as only 54% of the eligible couples in Bangladesh use modern contraception^28^, and this percentage is even lower for LAMC (33.54%), as found in this study. In Bangladesh, unintended conception and unplanned birth are very high, contributing 49% of all conceptions and 22% of all live births, respectively^28^. Many women who become pregnant unintentionally tend to go for the traditional procedure of abortion outside maternal healthcare services. All these issues lead to a higher number of maternal and child morbidity and mortality^9,36^. The findings of this current study indicate that these burdens can be overcome significantly by ensuring LAMC in the nearest health facilities and improving the service quality.

In Bangladesh, the programmes for improving health facility management, infrastructure and accessibility to LAMC were first taken into account as part of the seventh five-year plan (2016-2020)^37^— and are currently functional as part of the eighth five-year plan (2021-2026)^38^. However, no notable progress has been made to date. Providing contraception services, including LAMC, via family planning workers (FPW) (who visit couples’ homes fortnightly) is still the fundamental way to provide family planning services and contraception at the community level^28^. An advantage of this approach is that couples who are not interested in accessing services from health facilities can still access contraception services. However, this approach has two major disadvantages: i) FPW usually do not receive specialized medical education and ii) FPW are restricted to provide non-surgical contraception methods, mainly pills and condoms ^9,39^. Consequently, pills (25%) and condoms (7%) are the major contraceptives in Bangladesh, and are used by 60% of the overall contraception users^28^. This pattern is different from many other LMICs where condoms and female sterilization are the dominant contraception methods^40^. These indicate the dominant role of FPW in providing contraception services over health facilities ^9,31^. Furthermore, the failure rates of pills (7%) and condoms (13-21%) are very high at their typical use^41^. Therefore, the ongoing provision of contraception services via FPW needs to be improved to reduce unintended pregnancies and related adverse consequences^9^. Effective linkages between FPW and health facilities can be crucial in this regard.

There are many challenges to the increased use of LAMC in Bangladesh. FPWs play a major role in creating awareness about LAMC use among women in rural areas. This possibly explains a relatively high likelihood of LAMC use among disadvantaged groups, which is not consistent with the findings of other studies in LMICs and Bangladesh^1,17,18,42^. However, LAMC use is still relatively low, even among the women of the poorest and poorer wealth quintiles. LAMC use is unlikely to increase unless the coordination between FPWs and health facilities is improved. FPWs cannot provide LAMC services but can only refer couples to health facilities ^43^. FPWs work under the Directorate General of Family Planning Services and health facilities work under Directorate General of Health Services ^44^ and there is a lack of coordination due to differences in reporting arrangement and job descriptions ^9^. Although several programs to increase LAMC use have been taken in Bangladesh, including payment for sterilization or vasectomy, progress is unlikely to be significant unless the provider-level challenges are addressed ^45,46^.

Our findings suggest improvement in the management and infrastructure of health facilities has significant contributions to the increased use of LAMC. Healthcare facilities are overcrowded and are not congenial for women and healthcare providers to discuss sensitive topics such as LAMC use^9^. Also, there is a shortage of trained healthcare professionals assigned to LAMC and other contraception services^39,47^. Moreover, healthcare providers are expected to provide counselling in conjunction with LAMC ^38^. However, many practitioners, particularly those who completed Bachelor of Medicine or Bachelor of Surgery degree, may not prioritise counselling over other competing tasks. Moreover, in most healthcare facilities, a significant portion of healthcare providers are male. Women usually do not feel comfortable in accessing contraception services from them^39,47^. These challenges are even deep-rooted in rural health facilities along with poor management including inadequate medical supplies, inability to use modern technologies in providing health services and absence of assigned healthcare personnels^48^. Consequently, women in rural areas are less likely to use LAMC.

The availability of LAMC-providing health facilities in short distances is an important predictor of LAMC uptake. These observations are consistent with the findings from other LMICs such as Turkey and Zambia ^49-51^ and an indication that a portion of couples in Bangladesh do not access LAMC although they had an intention to use them. There may have a compound effect of this disadvantage for women of relatively low knowledge of LAMC. Social level barriers and taboo, a low agency of decision making and financial ability - all these can negatively affect the willingness to visit health facilities if they are located far away^9^. Together, this highlights the requirement of more healthcare facilities that can provide LAMC at the community level along with awareness-building about the benefits of LAMC use. Engaging 13,221 community clinics that are currently functioning could accelerate the initiative. Priority should be given in areas with a relatively low number of health facilities.

This study has several limitations. Data analysed in this study were extracted from cross-sectional surveys. Therefore, the findings are correlational only, not casual. To secure the privacy of the respondents, BDHS displaced clusters’ locations up 0-5 km for rural areas and 0-2 km for urban areas. Therefore, the calculated average distance of the nearest health facility could be slightly different from the actual distance. However, the BDHS ensured that the new perturbed locations fell within the designated administrative boundaries. Therefore errors from displacement are likely to be random and minimum. A previous study found that the effect of this variation is insignificant ^52^. Regardless of these limitations, to our knowledge, this is the first study in the context of LMICs that examined the influence of health facility-level factors adjusted with the individual-, household-, and community-level factors. Clusters of LAMC use and non-use were identified, and the factors associated with LAMC were determined using multilevel regressions with adjustment for a wide range of factors. Therefore, the finding of this study is likely to be precise and can be generalisable in countries with similar features to Bangladesh and may help in making evidence-based policies.

## Conclusion

During 2017-2018, around 33% of women used LAMC in Bangladesh. Hot spot analysis shows LAMC use was higher in some parts of Rangpur and Rajshahi divisions and lower in some locations of the Sylhet, Chattogram, Barishal and Khulna divisions. Health facility-level factors, including health facility infrastructure, management, and readiness to provide LAMC were strong predictors of LAMC use in Bangladesh and the effects of the predictors were substantial in rural than urban areas. Health facilities providing LAMC located at a closer distance were positively associated with increased uptake of LAMC. Health facilities should be strengthened to provide LAMC and more health facilities of such capacities are needed. The current provision of family planning services through the field-level workers also needs to be strengthened with strong coordination and referral linkages with the health facilities.

## Data Availability

The datasets used and analysed in this study are available from the Measure DHS website: https://dhsprogram.com/data/available-datasets.cfm

## Acknowledgement

The authors thank the MEASURE DHS for granting access to the 2011 and 2017/18 BDHS data.

## Authors’ contributions

MNK designed the study, performed the data analysis, and wrote the first draft of this manuscript. MMI and SA critically reviewed and edited all versions of this manuscript. All authors approved this final version of the manuscript.

## References

1. Cahill N, Sonneveldt E, Stover J, et al. Modern contraceptive use, unmet need, and demand satisfied among women of reproductive age who are married or in a union in the focus countries of the Family Planning 2020 initiative: a systematic analysis using the Family Planning Estimation Tool. The Lancet 2018; 391(10123): 870–82.

2. Family Planning. The Family Planning 2020 Commitment 2020.

3. The World Bank. Contraceptive Prevalence, any methods (% of women ages 15-49). 2021

4. Nations U. Contraceptive use by method 2019 New York, USA 2019.

5. Family Planning. FP2020: Catalyzing Collaboration 2017–2018. June, 2021 ed; 2020.

6. MacQuarrie K. Unmet need for family planning among young women: levels and trends: ICF International; 2014.

7. World Health Organization. Contraception 2015 ed. Geneva, Switzarland; 2015.

8. Assembly G. Sustainable development goals. SDGs Transform Our World 2015; 2030.

9. Khan MN, Harris M, Loxton D. Modern contraceptive use following an unplanned birth in bangladesh: an analysis of national survey data. International perspectives on sexual and reproductive health 2020; 46: 77–87.

10. Bearak J, Popinchalk A, Ganatra B, et al. Unintended pregnancy and abortion by income, region, and the legal status of abortion: estimates from a comprehensive model for 1990–2019. The Lancet Global Health 2020; 8(9): e1152–e61.

11. Goossens J, Van Den Branden Y, Van der Sluys L, et al. The prevalence of unplanned pregnancy ending in birth, associated factors, and health outcomes. Human Reproduction 2016: 1–13.

12. Darroch JE, Singh S, Nadeau J. Contraception: an investment in lives, health and development. Issues in brief (Alan Guttmacher Institute) 2008; (5): 1–4.

13. Shukla A, Kumar A, Mozumdar A, et al. Association between modern contraceptive use and child mortality in India: A calendar data analysis of the National Family Health Survey (2015-16). SSM-population health 2020; 11: 100588.

14. Saha UR, van Soest A. Infant death clustering in families: magnitude, causes, and the influence of better health services, Bangladesh 1982–2005. Population studies 2011; 65(3): 273–87.

15. Government of the People’s Republic of Bangladesh. The Sixth-Five Year Plan, 2011-2016.. Dhaka, Bangladesh, 2011.: General Economic Division, Planning Commission, Ministry of Planning, Government of the People’s Republic of Bangladesh,, 2001.

16. Blumenberg C, Hellwig F, Ewerling F, Barros AJ. Socio-demographic and economic inequalities in modern contraception in 11 low-and middle-income countries: an analysis of the PMA2020 surveys. Reproductive Health 2020; 17: 1–13.

17. Apanga PA, Kumbeni MT, Ayamga EA, Ulanja MB, Akparibo R. Prevalence and factors associated with modern contraceptive use among women of reproductive age in 20 African countries: a large population-based study. BMJ open 2020; 10(9): e041103.

18. Lasong J, Zhang Y, Gebremedhin SA, et al. Determinants of modern contraceptive use among married women of reproductive age: a cross-sectional study in rural Zambia. BMJ open 2020; 10(3): e030980.

19. Ahinkorah BO, Seidu A-A, Appiah F, et al. Individual and community-level factors associated with modern contraceptive use among adolescent girls and young women in Mali: a mixed effects multilevel analysis of the 2018 Mali demographic and health survey. Contraception and reproductive medicine 2020; 5(1): 1–12.

20. Tegegne TK, Chojenta C, Forder PM, Getachew T, Smith R, Loxton D. Spatial variations and associated factors of modern contraceptive use in Ethiopia: a spatial and multilevel analysis. BMJ open 2020; 10(10): e037532.

21. Hong R, Montana L, Mishra V. Family planning services quality as a determinant of use of IUD in Egypt. BMC Health services research 2006; 6(1): 1–8.

22. Wang W. How family planning supply and the service environment affect contraceptive use: findings from four East African countries: International Health and Development, ICF International; 2012.

23. Rose M, Abderrahim N, Stanton C, Helsel D. Maternity care. A comparative report on the availability and use of maternity services. Evaluation 1999.

24. Hossain M, Khan M, Ababneh F, Shaw J. Identifying factors influencing contraceptive use in Bangladesh: evidence from BDHS 2014 data. BMC public health 2018; 18(1): 1–14.

25. Haq I, Sakib S, Talukder A. Sociodemographic factors on contraceptive use among ever-married women of reproductive age: evidence from three demographic and health surveys in Bangladesh. Medical Sciences 2017; 5(4): 31.

26. Skiles MP, Burgert CR, Curtis SL, Spencer J. Geographically linking population and facility surveys: methodological considerations. Population health metrics 2013; 11(1): 1–13.

27. Household LD. DHS SPATIAL ANALYSIS REPORTS 10. 2014.

28. National Institute of Population Research and Training (NIPORT) aI. Bangladesh Demographic and Health Survey 2017-18 Dhaka, Bangladesh, and Rockville, Maryland, USA: NIPORT and ICF., 2020.

29. 2019 NIoPRaTNaI. Bangladesh Health Facility Survey 2017. Dhaka, Bangladesh: NIPORT, ACPR, and ICF., 2019.

30. Organization WH. Service availability and readiness assessment (SARA). World Health Organization Geneva, Switzerland 2015.

31. National Institute of Population Research and Training (NIPORT) and ICF. Bangladesh Health Facility Survey 2017. Dhaka, Bangladesh: NIPORT, ACPR, and ICF., 2019.

32. Diez-Roux AV. Multilevel analysis in public health research. Annual review of public health 2000; 21(1): 171–92.

33. UN General Assembly. Transforming our world : the 2030 Agenda for Sustainable Development. A/RES/70/1 2015.

34. Planning Commission: General Economic Division. Eighth Five Year Plan (FY2020-FY2025): Promoting Posperity and Fostering Inclusiveness Government of the People’s Republic of Bangladesh, 2021.

35. Planning Commission: General Economic Division. Seventh Five Year Plan (FY2016–FY2020): Accelerating Growth, Empowering Citizens: Government of the People’s Republic of Bangladesh, 2016.

36. Khan MN, Harris ML, Loxton D. Assessing the effect of pregnancy intention at conception on the continuum of care in maternal healthcare services use in Bangladesh: Evidence from a nationally representative cross-sectional survey. PloS one 2020; 15(11): e0242729.

37. Government of the People’s Republic of Bangladesh. 7th Five Year Plan, FY 2016-FY2020. General Economics Division, Planning Commission, Dhaka, Bangladesh., 2016.

38. Government of the People’s Republic of Bangladesh. 8th Five Year Plan, FY 2021-FY2026. General Economics Division, Planning Commission, Dhaka, Bangladesh., 2021.

39. Khan MN, Islam, M Mofizul,. Pregnancy resulting from contraceptive failure is increasing in Bangladesh: evidence from nationally representative demographic and health survey. Contraception (under-review) 2021.

40. Schuler SR, Hashemi SM, Jenkins AH. Bangladesh’s family planning success story: a gender perspective. International Family Planning Perspectives 1995: 132–66.

41. Trussell J, Guthrie K. Choosing a contraceptive: efficacy, safety, and personal considerations. Hatcher RA, Trussell J, Nelson AL, Cates W, Stewart FH, Kowal D Contraceptive technology 19th revised ed New York (NY): Ardent Media, Inc 2007: 19–47.

42. Huda FA, Robertson Y, Chowdhuri S, Sarker BK, Reichenbach L, Somrongthong R. Contraceptive practices among married women of reproductive age in Bangladesh: a review of the evidence. Reproductive health 2017; 14(1): 69-.

43. Das TR. Family planning program of Bangladesh: achievements and challenges. South East Asia Journal of Public Health 2016; 6(1): 1–2.

44. Organization WH. Bangladesh health system review: Manila: WHO Regional Office for the Western Pacific; 2015.

45. Donahoe DA. Men and family Planning in Bangladesh: A review of the literature: Population Council; 1996.

46. Cleland J, Mauldin WP. The promotion of family planning by financial payments: the case of Bangladesh. Studies in Family Planning 1991; 22(1): 1–18.

47. Khan MN, Islam, M Mofizul,. Women’s experience of unintended pregnancy and its effect on changes in pre-pregnancy contraceptive methods: Evidence from a nationally representative survey. Studies in Family Planning (under-review) 2021.

48. Hassan N, Rashid H, Das R. Smart Health Management System for Rural Area of Bangladesh Utilizing Smartphone and NID. Health care; 1: 3.

49. Silumbwe A, Nkole T, Munakampe MN, et al. Community and health systems barriers and enablers to family planning and contraceptive services provision and use in Kabwe District, Zambia. BMC Health Services Research 2018; 18(1): 390.

50. Sato R, Rohr J, Huber-Krum S, et al. Effect of distance to health facilities and access to contraceptive services among urban Turkish women. The European Journal of Contraception & Reproductive Health Care 2021: 1–9.

51. Wang W, Mallick L. Understanding the relationship between family planning method choices and modern contraceptive use: an analysis of geographically linked population and health facilities data in Haiti. BMJ Global Health 2019; 4(Suppl 5): e000765.

52. Sato R KM, Elewonibi B, Mhande M, Msuya S, Shah I, Canning D,. Distance to Facility and Healthcare Utilization in Tanzania: Unbiased and Consistent Estimation using Perturbed Location Data. Population Association of America New York, USA Population Association of America 2019.

